# Methodological approaches to optimize multiplex oral fluid SARS-CoV-2 IgG assay performance and correlation with serologic and neutralizing antibody responses

**DOI:** 10.1101/2022.12.22.22283858

**Authors:** Nora Pisanic, Annukka A. R. Antar, Kate Kruczynski, Magdielis Gregory Rivera, Santosh Dhakal, Kristoffer Spicer, Pranay R. Randad, Andrew Pekosz, Sabra L. Klein, Michael J. Betenbaugh, Barbara Detrick, William Clarke, David L. Thomas, Yukari C. Manabe, Christopher D. Heaney

## Abstract

**Background:** Oral fluid (hereafter, saliva) is a non-invasive and attractive alternative to blood for SARS-CoV-2 IgG testing; however, the heterogeneity of saliva as a matrix poses challenges for immunoassay performance.

**Objectives:** To optimize performance of a magnetic microparticle-based multiplex immunoassay (MIA) for SARS-CoV-2 IgG measurement in saliva, with consideration of: i) threshold setting and validation across different MIA bead batches; ii) sample qualification based on salivary total IgG concentration; iii) calibration to U.S. SARS-CoV-2 serological standard binding antibody units (BAU); and iv) correlations with blood-based SARS-CoV-2 serological and neutralizing antibody (nAb) assays.

**Methods:** The salivary SARS-CoV-2 IgG MIA included 2 nucleocapsid (N), 3 receptor-binding domain (RBD), and 2 spike protein (S) antigens. Gingival crevicular fluid (GCF) swab saliva samples were collected before December, 2019 (n=555) and after molecular test-confirmed SARS-CoV-2 infection from 113 individuals (providing up to 5 repeated-measures; n=398) and used to optimize and validate MIA performance (total n=953). Combinations of IgG responses to N, RBD and S and total salivary IgG concentration (μg/mL) as a qualifier of nonreactive samples were optimized and validated, calibrated to the U.S. SARS-CoV-2 serological standard, and correlated with blood-based SARS-CoV-2 IgG ELISA and nAb assays.

**Results:** The sum of signal to cutoff (S/Co) to all seven MIA SARS-CoV-2 antigens and disqualification of nonreactive saliva samples with ≤15 μg/mL total IgG led to correct classification of 62/62 positives (sensitivity [Se]=100.0%; 95% confidence interval [CI]=94.8%, 100.0%) and 108/109 negatives (specificity [Sp]=99.1%; 95% CI=97.3%, 100.0%) at 8-million beads coupling scale and 80/81 positives (Se=98.8%; 95% CI=93.3%, 100.0%] and 127/127 negatives (Sp=100%; 95% CI=97.1%, 100.0%) at 20-million beads coupling scale. Salivary SARS-CoV-2 IgG crossed the MIA cutoff of 0.1 BAU/mL on average 9 days post-COVID-19 symptom onset and peaked around day 30. Among n=30 matched saliva and plasma samples, salivary SARS-CoV-2 MIA IgG levels correlated with corresponding-antigen plasma ELISA IgG (N: ρ=0.67, RBD: ρ=0.76, S: ρ=0.82; all *p*<0.0001). Correlations of plasma SARS-CoV-2 nAb assay area under the curve (AUC) with salivary MIA IgG (N: ρ=0.68, RBD: ρ=0.78, S: ρ=0.79; all *p*<0.0001) and with plasma ELISA IgG (N: ρ=0.76, RBD: ρ=0.79, S: ρ=0.76; *p*<0.0001) were similar.

**Conclusions:** A salivary SARS-CoV-2 IgG MIA produced consistently high Se (>98.8%) and Sp (>99.1%) across two bead coupling scales and correlations with nAb responses that were similar to blood-based SARS-CoV-2 IgG ELISA data. This non-invasive salivary SARS-CoV-2 IgG MIA could increase engagement of vulnerable populations and improve broad understanding of humoral immunity (kinetics and gaps) within the evolving context of booster vaccination, viral variants and waning immunity.

## 1. INTRODUCTION

Monitoring SARS-CoV-2 antibody responses^1,2^ induced by natural infection and/or vaccination is important to estimate seroprevalence and to understand transmission patterns that can be driven by asymptomatic exposure or infection, waning immunity, and gaps in humoral immunity^3–5^. Most SARS-CoV-2 seroprevalence studies utilize peripheral blood-based serologic assays, which require phlebotomy or needle stick (e.g., dried blood spots). Peripheral blood collection is typically justified by the high performance of blood-based SARS-CoV-2 serologic assays and their correlation with SARS-CoV-2 neutralizing antibody (nAb) activity.^6–8^ However, participant burdens associated with phlebotomy makes it unsuitable for repeated, routine antibody testing. Further, seroprevalence studies requiring participants to complete in-person visits at a clinical site for study procedures and phlebotomy tend to miss under-represented groups, such as under-resourced communities, populations with limited access to healthcare, populations who mistrust the medical establishment, and people who fear needle sticks.^9,10^ To overcome these challenges and increase opportunity for broad population-scale estimates of SARS-CoV-2 seroprevalence, several groups^11–13^ have developed SARS-CoV-2 antibody tests using saliva and have shown that saliva-based antibody tests can be reasonably accurate; however, none approach the highly accurate test performance of the best blood-based SARS-CoV-2 antibody tests. Most achieve high specificity (e.g., >99%), but fail to achieve sensitivity >90-95%.^14,15^

We recently developed a multiplex SARS-CoV-2 antibody assay for use with self-collected saliva.^16^ Saliva for this assay is collected using a lollipop-like sponge on a stick, with which participants were instructed to brush the line between their teeth and gums for 1-2 minutes, in a motion similar to brushing their teeth. This collection method stimulates leakage of antibody-enriched serum-derived gingival crevicular fluid (GCF)^17^ from between the gums and teeth that is then absorbed by the swab. Saliva can be collected at home and mailed to a centralized lab for testing of antibodies to SARS-CoV-2 nucleocapsid (N), receptor-binding domain (RBD) and spike (S) proteins with an in-house microparticle bead-based multiplex immunoassay (MIA) based on Luminex xMAP technology.^4,18,19^

Here we aimed to optimize the SARS-CoV-2 IgG multiplex assay for high accuracy, to characterize and validate the assay using pre-COVID-19 era saliva samples and prospectively collected saliva samples from study participants who tested positive for SARS-CoV-2 by RT-PCR. We coupled two MIA bead batches at different scales and determined assay sensitivity and specificity across bead batches using close to 1,000 saliva samples. We investigated how sample qualification based on the total salivary IgG concentration affects SARS-CoV-2 IgG assay performance and kinetic trajectories post COVID-19 symptoms onset. We estimated time to SARS-CoV-2 IgG immune-conversion and time to establish a robust IgG level and investigated differences in anti-N, -RBD and -S IgG kinetics post-COVID-19. We correlated salivary IgG levels with blood-based IgG levels (anti-N, -RBD and -S IgG ELISA) and with plasma SARS-CoV-2 nAb assay area under the curve (AUC). And lastly, motivated by the goal to achieve comparability with other quantitative serological assays, we calibrated our quantitative assay standards to the U.S. SARS-CoV-2 serological standard to estimate the concentration of salivary SARS-CoV-2 IgG in samples from the pre-COVID-19 era and post SARS-CoV-2 infection.

## 2. METHODS

### 2.1. Study population

Samples were collected longitudinally from consented research participants who had tested positive for SARS-CoV-2 by RT-PCR in the 1-3 days prior to enrollment and before vaccines were available (April – September 2020).^18,20^Archived saliva samples that had been self-collected with Oracol swabs as part of different research studies before December 2019 were used as presumed negative samples.^21–23^

### 2.2. Saliva self-collection and sample processing

Upon enrollment, each participant received a self-testing kit containing saliva collection swabs (Oracol S14, Malvern Medical Developments, Worcester, UK) and other study supplies by courier delivery. After receipt of the testing supplies a study coordinator scheduled a video or phone visit (day 0) to instruct the participant on saliva self-collection procedures, collect basic demographics, and note the date of COVID-19 symptom onset. Follow-up phone or video visits, during which participants self-collected additional saliva samples, were scheduled on study days 3, 7, and 14. On study day 28, participants were invited for an in-person visit during which blood and saliva were collected. Study staff collected blood by venipuncture into EDTA tubes and the resulting plasma was separated from solid blood components through centrifugation. Self-collected saliva from study days 0, 3, and 7 were stored in a freezer (−20°C) by the participant after collection until the last sample was collected at home on day 14. Swabs were then returned to the study by courier in a package with ice packs. Swabs collected during the day 28 in-person clinical follow-up visit were transferred directly to the processing lab. Upon receipt at the lab, swabs were centrifuged at 1,500 g for 10 minutes to separate saliva from the sponge and then heat-inactivated at 56°C for 1 hour in a water bath to inactivate SARS-CoV-2 virus if present. Samples were stored at ≤-20°C prior to testing. Presumed SARS-CoV-2 IgG negative archived saliva samples were also heat-inactivated at 56°C for 1 hour prior to testing.

### 2.3. Coupling of antigens and antibodies to magnetic beads

Saliva samples were tested using a modified version of a previously described multiplex SARS-CoV-2 immunoassay^16^ based on Luminex xMAP technology. The modified multiplex assay version was composed of 20 unique magnetic bead sets (MagPlex microspheres), instead of 12 sets. Each set was coupled covalently with antigen, antibody or BSA (controls), as described previously.^16^ The assay included SARS-CoV-2 nucleocapsid (N), receptor binding domain (RBD), and spike (S) antigens (all Wuhan 2019 strain), SARS, MERS, RSV, human coronavirus 229E, NL63, HKU1, and OC43 antigens in addition to control antibodies and proteins (BSA, anti-human IgG, IgM, IgA antibody; see **Table S1**). Two different multiplex assay lots, one at 8 million beads coupling scale (sufficient for approximately 80 assay plates) and one at 20 million beads coupling scale (for ∼200 assay plates), were produced and their performance compared.

### 2.4. Salivary SARS-CoV-2 IgG multiplex immunoassay (MIA)

Saliva was tested for IgG binding to SARS-CoV-2 and additional virus antigens in the MIA as described previously.^16^ Briefly, saliva was thawed and centrifuged for 5 minutes at 20,000 g. Then 10 μL saliva supernatant was added to a 96-well microtiter plate containing 40 μL PBST with 1% BSA (assay buffer) and 1,000 coupled beads per bead set in each well. Each plate contained 1-2 blank wells with assay buffer instead of sample that were used for background fluorescence subtraction. Positive controls were created by spiking SARS-CoV-2 IgG positive saliva with high IgG levels to SARS-CoV-2 antigens into pre-pandemic negative saliva. Pre-pandemic saliva was used as negative control. Phycoerythrin-labeled anti-human IgG diluted 1:100 in assay buffer was used to detect the IgG signal in saliva (see **Table S1**). Assay plates were read on a Luminex MAGPIX instrument.

After assay optimization, an 8-point standard curve of 3-fold serial dilutions was included on each plate in duplicate. The standard consisted of plasma from a PCR-confirmed COVID-19 study participant with high SARS-CoV-2 IgG levels diluted in assay buffer. Arbitrary units (AU) ranging from 10,000 AU for the highest standard to ∼5 AU for the lowest standard were assigned to each standard. The IgG concentration to SARS-CoV-2 antigens in AU was calculated with xPONENT Software (Luminex) using a weighted 5-parameter logistic curve fit. The assay precision within and between assay plates was determined by assessing replicate measurements of contrived saliva high and low positive controls and a lower limit of quantification control; ten random saliva samples collected prior to the COVID-19 pandemic were tested, percent recovery of a spiked and serially diluted saliva sample was assessed, and precision around the assay cutoff was assessed with contrived saliva samples near (+/-25% and +/-50%) and at the cutoff measured in replicate. Last, the assay performance was validated using pre-COVID-19 samples (presumed negatives) and samples collected from COVID-19 study participants (confirmed by FDA-EUA RT-PCR test).

### 2.5. Salivary SARS-CoV-2 MIA calibration to the U.S. national SARS-CoV-2 serology standard

A dilution series of the U.S. national SARS-CoV-2 serology standard (8 points in duplicate) was included as unknowns to measure its concentration in AUs. Likewise, a dilution series of our in-house standard was included in the SARS-CoV-2 IgG multiplex assay as unknowns using dilutions of the national serology standard as the assay standard to estimate the in-house standard’s concentration in U.S. serology standard units (BAU/mL) to determine the concentration equivalent of the assay cutoff in BAU/mL.

### 2.6. Salivary total IgG ELISA

The total IgG concentration in saliva was determined using Salimetrics Salivary Human Total IgG ELISA Kits according to the manufacturer’s instructions with two modifications. The incubation times with diluted saliva sample and with detect antibody were reduced to 1 hour each instead of 2 hours. This modification has been discussed with and approved by the manufacturer. ELISA standards and high and low total salivary IgG assay controls provided with the kit with defined total IgG concentration ranges were assessed on each plate.

### 2.7. Statistical analysis

The blank-subtracted (net) median fluorescence intensity (MFI) was used for data analysis and for standard curve fitting. IgG binding expressed in AU and in MFI were compared. Cutoffs to discriminate SARS-CoV-2 IgG positive from negative samples for each individual SARS-CoV-2 antigen were defined as the average net MFI (or AU) plus three standard deviations (SD) of samples collected prior to December 2019 (pre-COVID-19 negatives). The resulting sensitivity and specificity of individual SARS-CoV-2 antigens were calculated using saliva from COVID-19 study participants (positives) and pre-COVID-19 saliva samples (negatives).

In addition to relying on individual antigens to determine presence of SARS-CoV-2 IgG, several algorithms involving IgG binding to multiple antigens were explored. Algorithms tested included positive classification if the IgG signal is above the cutoff (signal to cutoff [S/Co]>1) for 1) at least one nucleocapsid and one RBD or S antigen; 2) two or more RBD or spike (S) antigens; or 3) positive for either algorithm (1) or (2). Additional algorithms included the sum of IgG S/Cο to multiple antigens with the cutoffs for such combinations calculated correspondingly, i.e., the mean plus 3 SDs of each algorithm tested in pre-pandemic negatives. We determined assay performance based on 4) anti-N IgG; 5) anti-RBD IgG; 6) anti-S IgG binding; and 7) IgG binding to all seven N, RBD and S antigens combined. Recognizing that samples collected post COVID-19 vaccination will contain anti-RBD and anti-S IgG, we also determined the sensitivity to classify prior SARS-CoV-2 infection correctly by combining algorithms (7) and (4). Algorithm 7 is based on IgG binding to any combination of antigens and would thus classify post-vaccine samples as positive, whereas algorithm 4 tests for presence of anti-N IgG, which should only be present post natural infection but not post vaccination. Lastly, we also explored whether normalizing the SARS-CoV-2 IgG signal with the total salivary IgG (tIgG) concentration in μg/mL would improve the assay performance; 8) sum S/Co to N/RBD/S divided tIgG.

Next, we evaluated the influence of total salivary IgG on assay performance. Three minimum total IgG concentrations, 5 μg/mL, 10 μg/mL and 15 μg/mL, to “qualify” algorithm negative samples were explored. The minimum total IgG requirement quality control (QC) measure was only applied when samples tested negative using the applied algorithm, i.e., samples testing positive for SARS-CoV-2 IgG were not subject to the minimum total IgG concentration requirement, whereas samples that tested negative failed QC (insufficient total IgG for correct classification) if the total IgG concentration was below the minimum concentration defined. The number of samples that failed QC was also calculated.

Spearman correlation coefficients were calculated to estimate the correlation between plasma neutralizing SARS-CoV-2 antibody area under the curve (AUC)^8^, plasma anti-N, RBD and S IgG AUC measured by ELISA^8^ and corresponding salivary SARS-CoV-2 IgG. Loess regression was used to visualize IgG antibody kinetics to SARS-CoV-2 in the multiplex test after COVID-19 symptoms onset. Mononuclear growth models were used to calculate time from COVID-19 symptoms onset to seroconversion and to reach maximum IgG levels. Means, standard deviations and %CV were calculated to characterize within and between assay plate precision and precision near the assay cutoff. To determine assay performance, samples were classified as positive, negative, or indeterminate and sensitivity, specificity and overall concordance were calculated. Statistical analyses were performed in SAS University Edition Release 3.8, SAS Institute Inc., Cary, NC, USA, and with RStudio Version 1.4.

## 3. RESULTS

### 3.1. Saliva samples used for SARS-CoV-2 IgG MIA characterization and optimization

Between April and September 2020, 113 ambulatory COVID-19 study participants provided 398 saliva samples. Most samples (66%) were collected more than two weeks after COVID-19 symptoms started. About two thirds of participants (n=75; 67%) provided at least 2 samples and 50 (45%) provided saliva samples for all five study events, i.e., one enrollment sample and follow-up samples on study days 3, 7, 14 and 28. Approximately three-quarters of these samples (n=293; 74%) were used to characterize and optimize the multiplex assay and the remaining quarter (n=105; 26%) were used for assay validation. Archived saliva samples collected before December 2019 (n=555) were used as negative controls. Correspondingly, a larger set of pre-COVID-19-era samples was used for assay optimization (n=362; 65%) and a smaller set for validation (n=193; 35%; **Table 1**).

**Table 1.** Self-collected COVID-19 study participant saliva samples and pre-COVID-19 era archived saliva samples for SARS-CoV-2 IgG multiplex test optimization and validation.

| Testing Phase | COVID-19 study participant saliva samples |  |  | Pre-COVID-19 saliva samples | Total |
| --- | --- | --- | --- | --- | --- |
|  | Days after COVID-19 symptoms onset |  |  |  |  |
|  | 1-7 days | 8-14 days | >14 days |  |  |
| Threshold Setting, n (%) | 41 (6%)<sup>#</sup> | 78 (12%)<sup>#</sup> | 174 (27%)<sup>#</sup> | 362 (55%)<sup>#</sup> | 655 (69%)* |
| Validation, n (%) | 4 (1%)<sup>#</sup> | 14 (5%)<sup>#</sup> | 87 (29%)<sup>#</sup> | 193 (65%)<sup>#</sup> | 298 (31%)* |
| Total, n (%) | 45 (5%)<sup>#</sup> | 92 (10%)<sup>#</sup> | 261 (27%)<sup>#</sup> | 555 (58%)<sup>#</sup> | 953 (100%) |

<sup>#</sup>Distribution within testing phase/row; \*Distribution between threshold setting and validation phase

### 3.2. Threshold setting

#### 3.2.1. Performance of individual antigens in SARS-CoV-2 IgG MIA

The sensitivity and specificity for each individual SARS-CoV-2 antigen were calculated as a first step to characterize each antigen’s performance classifying presumed SARS-CoV-IgG positive samples from COVID-19 study participants and presumed negative saliva samples collected prior to the COVID-19 pandemic correctly. In accordance with FDA-guidelines for antibody testing, only samples collected at least two weeks after COVID-19 symptoms onset were used to determine sensitivity.^24^ In general, higher sensitivity was achieved at the cost of lower specificity and vice versa (**Table S2**). For example, the Sino Biological RBD antigen results in 84% sensitivity but only 98% specificity, whereas GenScript’s RBD antigen results in higher specificity (99%) but lower sensitivity (78%). With the exception of one S antigen (1% sensitivity) and a SARS N antigen, which showed equivalent performance to GenScript N and was derived from a SARS 2002, not SARS-CoV-2 strain, we concluded that the remaining antigens could contribute to a combined algorithm for sample classification.

#### 3.2.2. Algorithms to improve salivary SARS-CoV-2 IgG MIA performance

The assay performance was calculated using eight algorithms that combined IgG binding to two or more of the seven SARS-CoV-2 antigens (**Table S3**). Algorithms included positive classification if the IgG signal is above the cutoff for 1) ≥1 N and ≥1 RBD or S antigen; 2) ≥2 RBD or S antigens; 3) positive for either algorithm (1) or (2). Additional classifications were based on the sum of IgG S/Cο to the SARS-CoV-2 antigens represented in the assay with the cutoffs for such combinations calculated correspondingly (mean plus 3 SDs in pre-pandemic saliva samples): 4) N (n=2); 5) RBD (n=3); 6) S (n=2); and 7) IgG binding to all seven N, RBD and S antigens combined. Recognizing that samples collected post COVID-19 vaccination will contain anti-RBD and anti-S IgG, we also determined the sensitivity to classify prior SARS-CoV-2 infection correctly by combining algorithms (7) and (4). Algorithm 7 is based on IgG binding to any combination of antigens and would thus classify post-vaccine samples as positive, whereas algorithm 4 tests for presence of anti-N IgG, which should only be present post natural infection, not post vaccination. Lastly, we also explored whether normalizing the SARS-CoV-2 IgG signal with the total salivary IgG (tIgG) concentration in μg/mL would improve the assay performance; 8) sum S/Co to N/RBD/S divided tIgG. Most combinations improved assay performance compared to relying on any single antigen for sample classification. The assay sensitivity increased to 84.5% with algorithm 7 that combines the IgG response to seven antigens while maintaining high specificity (98.6%). Algorithm 3 (**Table S3**, positive if IgG signal above cutoff for at least one N and one RBD/S antigen or two or more RBD/S antigens) also improved overall test accuracy. Normalizing the SARS-CoV-2 IgG response with the total IgG concentration (Algorithm 8, **Table S3**) yielded poor sensitivity (<50%, **Table S3**).

#### 3.2.3. Effect of total salivary IgG concentration on SARS-CoV-2 IgG MIA performance

Large differences in total antibody concentration between saliva samples compared to the relatively small concentration difference between blood samples need to be overcome for salivary SARS-CoV-2 IgG tests to perform equivalently to a typical blood-based SARS-CoV-2 antibody test. We therefore investigated differences in total IgG concentration between presumably false-negative saliva samples and saliva samples that were classified correctly as SARS-CoV-2 IgG positive using the best performing algorithm, algorithm 7, the sum of SARS-CoV-2 IgG S/Co to N, RBD and S antigens.

Total salivary IgG in samples from COVID-19 study participants that tested false negative for SARS-CoV-2 IgG was significantly lower than in samples classified correctly as positive (*p*<0.001; **Figure 1A**Error! Reference source not found.). However, many samples with low total IgG (e.g., <10 μg/mL) nonetheless contained enough SARS-CoV-2 IgG to cross the cutoff, likely because the proportion of SARS-CoV-2-specific IgG to total IgG was particularly high in such samples. We next examined the effect of total salivary IgG on the SARS-CoV-2 IgG signal in the first week, second week and more than two weeks post symptoms onset and in pre-COVID-19 samples. For this, saliva samples were divided into four categories of total IgG concentration in 5 μg/mL increments. SARS-CoV-2 IgG levels increased in each group with increasing total salivary IgG (**Figure 1B**). The difference in SARS-CoV-2 signal between archived pre-COVID-19 era samples and COVID-19 study samples collected >14 days post symptom onset was highest in the >10 μg/mL and 15 μg/mL total IgG categories. However, note that only pre-COVID-19 era samples in the highest category (>15 μg/mL) crossed the cutoff (false-positives).

**Figure 1.**
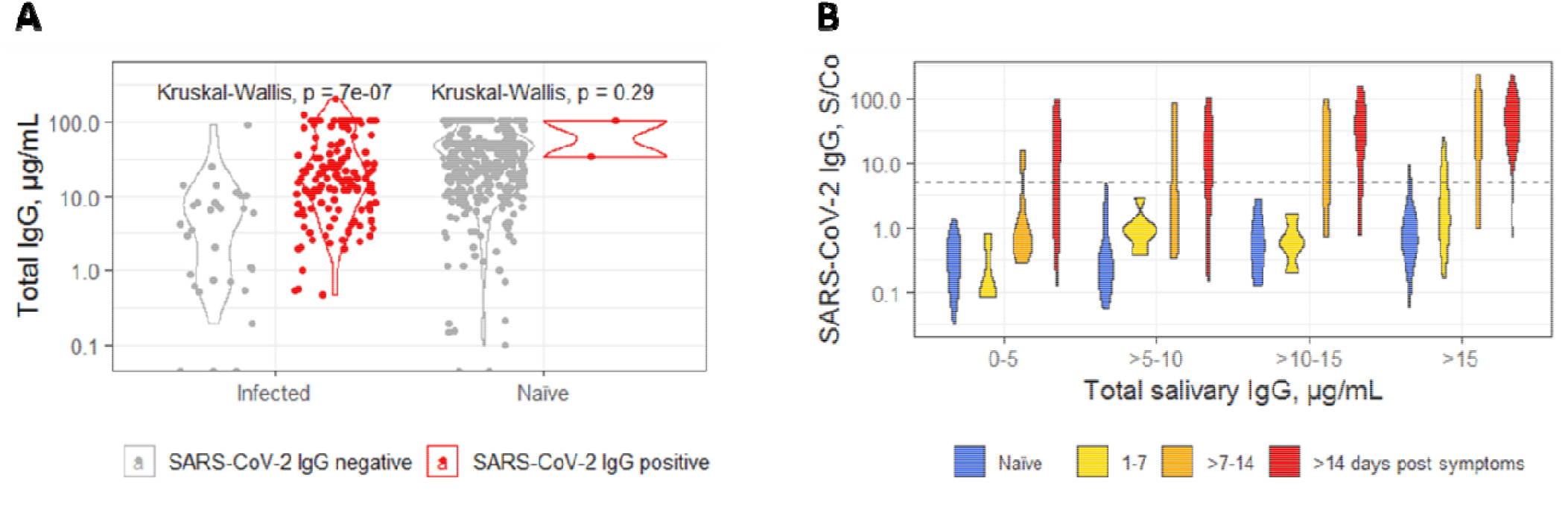
**A**. Distribution of salivary total IgG concentration in samples from COVID-19 study participants (Infected) and in archived pre-COVID-19 era samples (Naïve) stratified by SARS-CoV-2 IgG test outcome. **B**. SARS-CoV-2 IgG in pre-COVID-19 era and in samples collected 1-7 days, >7-14 days and >14 days post COVID-19 symptoms onset stratified by total IgG concentration. Note: SARS-CoV-2 IgG outcome was determined using the sum of IgG signal to cutoff (S/Co) to 2 N, 3 RBD and 2 S antigens (Table 2). Dotted line: cutoff.

#### 3.2.4. Establishment of a minimum total salivary IgG concentration as sample qualifier

The large proportion of samples with very low total IgG that tested false-negative for SARS-CoV-2 IgG led us to examine whether the disqualification of samples with insufficient total IgG could improve test accuracy. We tested this by defining a minimum total IgG concentration for samples to qualify as true negatives. This minimum total IgG qualification was only applied to samples that tested SARS-CoV-2 IgG negative using one of the algorithms. Samples with a positive classification were retained regardless of their total IgG concentration. We assessed the assay performance after defining minimum total IgG concentrations as sample qualifier in 5 μg/mL increments (5 μg/mL, 10 μg/mL and 15 μg/mL total salivary IgG). Pre-COVID-19 samples did not contribute to the cutoffs (mean plus 3 SDs of pre-COVID-19 samples) if the total IgG concentration was below the qualifying concentration. **Table 2** summarizes the multiplex assay performance for each minimum total IgG concentration chosen using the algorithms described above.

**Table 2.**
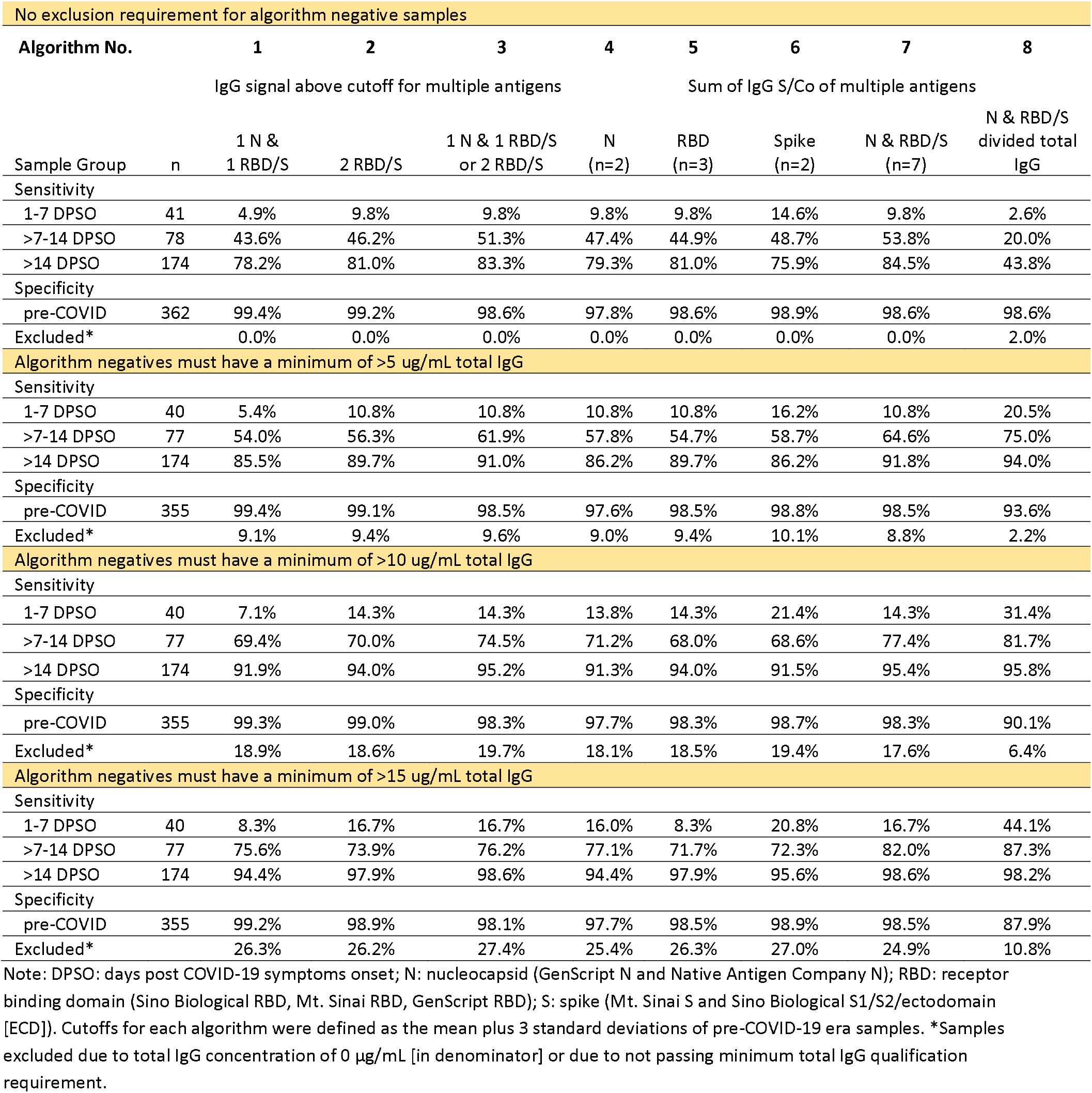
Effect of total IgG qualification of algorithm negative samples on assay performance.

**Table 3.**
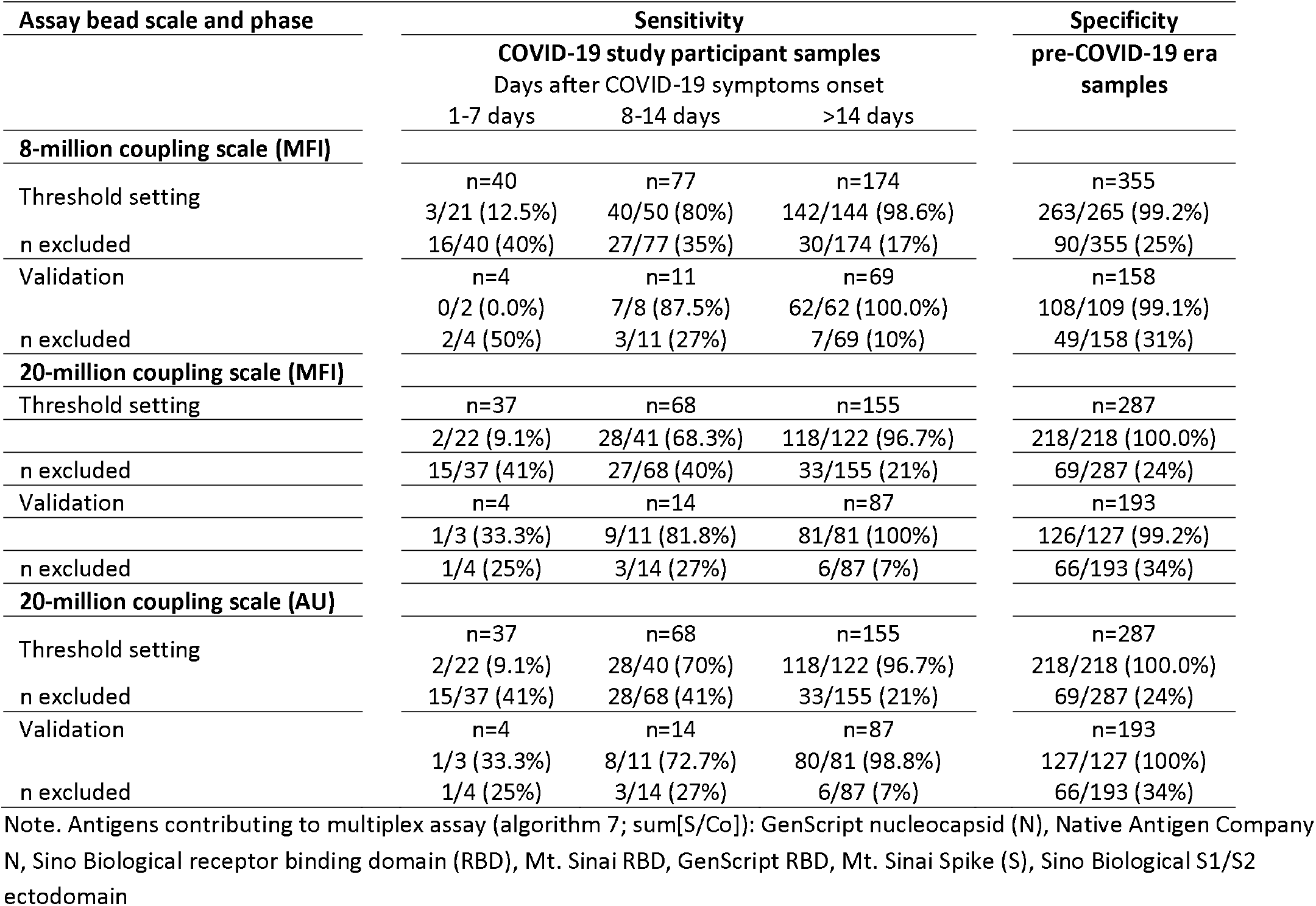
Salivary SARS-CoV-2 IgG MIA performance during threshold setting and validation phases.

| Assay bead scale and phase | Sensitivity |  |  | Specificity |
| --- | --- | --- | --- | --- |
|  | COVID-19 study participant samples |  |  | pre-COVID-19 era samples |
|  | Days after COVID-19 symptoms onset |  |  |  |
|  | 1-7 days | 8-14 days | >14 days |  |
| **8-million coupling scale (MFI)** |  |  |  |  |
| Threshold setting | n=40 | n=77 | n=174 | n=355 |
|  | 3/21 (12.5%) | 40/50 (80%) | 142/144 (98.6%) | 263/265 (99.2%) |
| n excluded | 16/40 (40%) | 27/77 (35%) | 30/174 (17%) | 90/355 (25%) |
| Validation | n=4 | n=11 | n=69 | n=158 |
|  | 0/2 (0.0%) | 7/8 (87.5%) | 62/62 (100.0%) | 108/109 (99.1%) |
| n excluded | 2/4 (50%) | 3/11 (27%) | 7/69 (10%) | 49/158 (31%) |
| **20-million coupling scale (MFI)** |  |  |  |  |
| Threshold setting | n=37 | n=68 | n=155 | n=287 |
|  | 2/22 (9.1%) | 28/41 (68.3%) | 118/122 (96.7%) | 218/218 (100.0%) |
| n excluded | 15/37 (41%) | 27/68 (40%) | 33/155 (21%) | 69/287 (24%) |
| Validation | n=4 | n=14 | n=87 | n=193 |
|  | 1/3 (33.3%) | 9/11 (81.8%) | 81/81 (100%) | 126/127 (99.2%) |
| n excluded | 1/4 (25%) | 3/14 (27%) | 6/87 (7%) | 66/193 (34%) |
| **20-million coupling scale (AU)** |  |  |  |  |
| Threshold setting | n=37 | n=68 | n=155 | n=287 |
|  | 2/22 (9.1%) | 28/40 (70%) | 118/122 (96.7%) | 218/218 (100.0%) |
| n excluded | 15/37 (41%) | 28/68 (41%) | 33/155 (21%) | 69/287 (24%) |
| Validation | n=4 | n=14 | n=87 | n=193 |
|  | 1/3 (33.3%) | 8/11 (72.7%) | 80/81 (98.8%) | 127/127 (100%) |
| n excluded | 1/4 (25%) | 3/14 (27%) | 6/87 (7%) | 66/193 (34%) |

Note. Antigens contributing to multiplex assay (algorithm 7; sum[S/Co]): GenScript nucleocapsid (N), Native Antigen Company N, Sino Biological receptor binding domain (RBD), Mt. Sinai RBD, GenScript RBD, Mt. Sinai Spike (S), Sino Biological S1/S2 ectodomain

The MIA performance improved with every 5 μg/mL increment in the minimum “required” total salivary IgG concentration to qualify negative samples (see **Table 2**). Note that the sum of S/Co to all seven SARS-CoV-2 IgG antigens (N & RBD/S, algorithm 7) resulted almost always in the highest test accuracy regardless of the minimum total salivary IgG concentration set forward to disqualify potentially false-negatives. We chose to move forward requiring SARS-CoV-2 IgG negative samples to contain a minimum IgG concentration of 15 μg/mL. This requirement did not apply to samples that were classified as positive. However, this minimum total IgG requirement also led to a loss or exclusion of samples for which no result could be determined (indeterminates). The sensitivity of the algorithm that “normalizes” the SARS-CoV-2 IgG signal with total IgG (algorithm 8) also improved significantly from <50% to >98% but at the cost of lower specificity (88%). This algorithm results, however, in the lowest proportion of samples excluded.

Using the sum of S/Co to all seven SARS-CoV-2 antigens (algorithm 7), **Error! Reference source not found**.A demonstrates how the minimum total IgG concentration applied to qualify samples as true SARS-CoV-2 IgG negatives improves the assay sensitivity from 84.5% without total IgG qualification to 98.6% with a minimum concentration of 15 μg/mL total IgG required. The proportion of indeterminate samples was lowest in the >14 days post symptom onset category (28/174 indeterminate samples; 16% at 15 μg/mL) and 25% of samples resulted in an indeterminate classification across the four groups.

#### 3.2.5. Optimization of salivary SARS-CoV-2 IgG MIA cutoff

To maximize assay accuracy, we defined 15 μg/mL as the minimum total IgG concentration required to qualify samples that tested SARS-CoV-2 negative as true negatives (**Figure 2**; **Figure 3A**). Next, we examined how fine-tuning the cutoff, rather than relying on the mean + 3 SD of pre-pandemic samples, influences assay performance and sample loss due to exclusion (Error! Reference source not found.**B**). Increasing the cutoff results in improved specificity (98.5% at a cutoff of 5 and 99.6% at a cutoff of 8 to 10), however, sensitivity decreased at a cutoff of 9 and higher (98.6% at cutoff 5 to 8; 97.8% at cutoff 9 and 95.6% at cutoff 10). The proportion of indeterminate samples increased from 22% at a cutoff of 5 to 24% at a cutoff of 10. The corresponding analysis and resulting assay performance of the two different bead batch coupling scales was remarkably similar. We also compared the assay performance using either the net fluorescence signal intensity (MFI) or standardized concentrations (AU) derived from the SARS-CoV-2 IgG standard curve, which was nearly identical (**Figures S1 and S2**).

**Figure 2:**
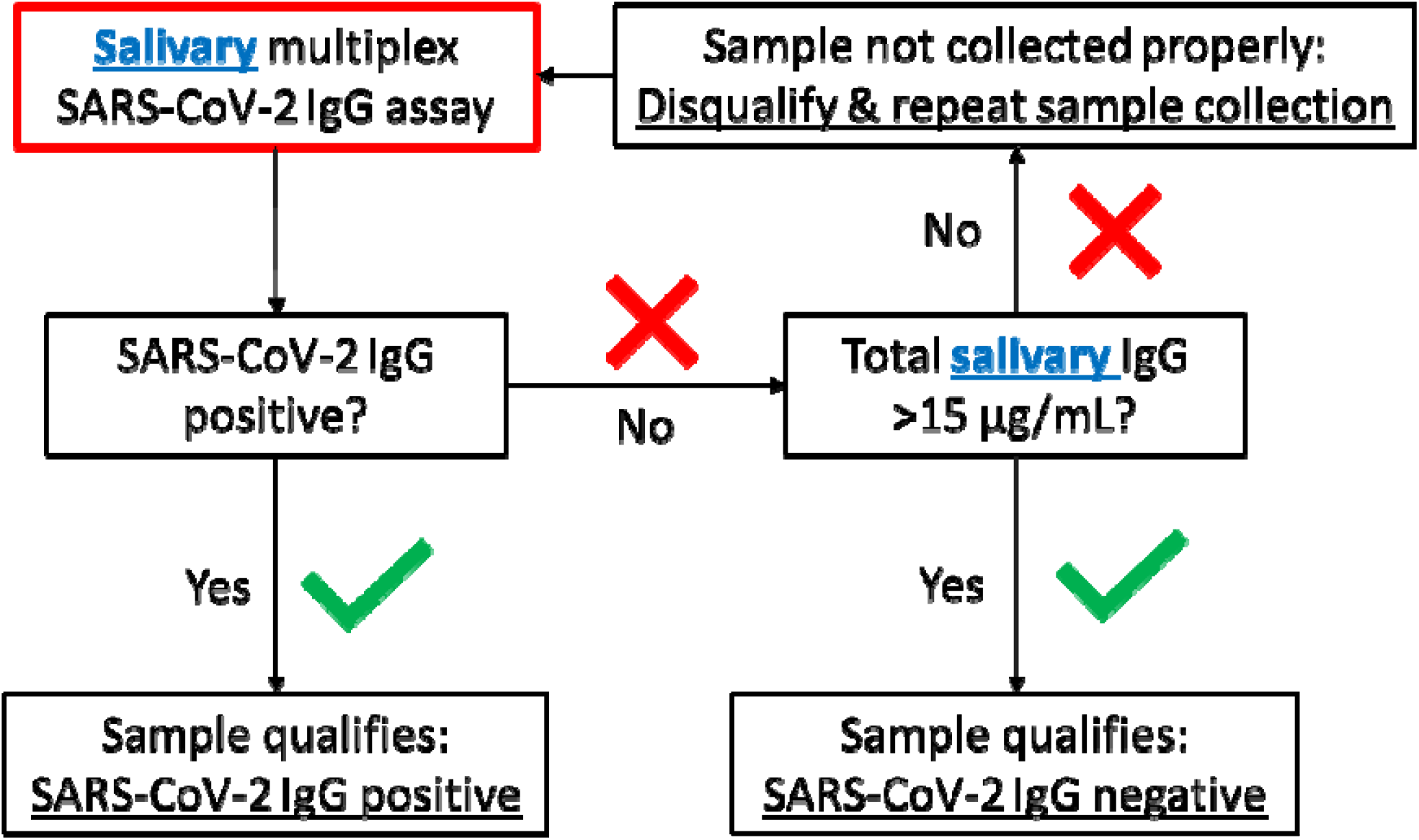
Sequence for salivary SARS-CoV-2 IgG MIA testing and qualification.

**Figure 3.**
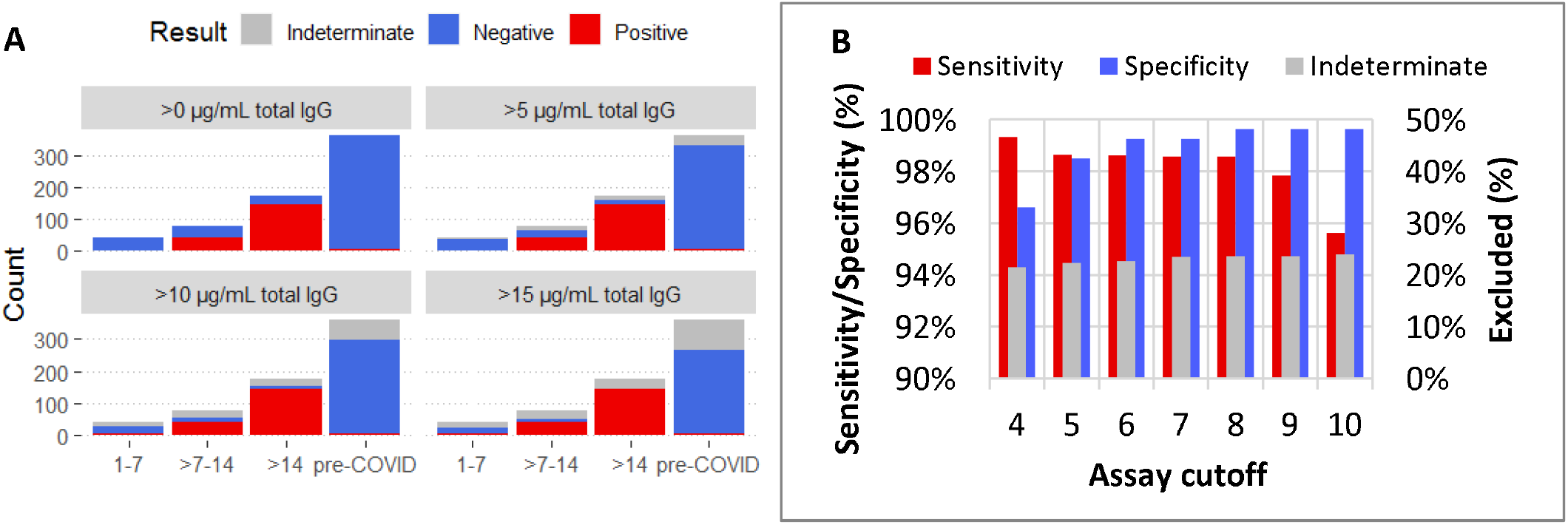
**A)** Effect of establishing a minimum total IgG concentration threshold for saliva qualification on MIA performance**; B)** MIA performance and sample exclusion at various cutoffs (8 million beads coupling scale).

### 3.3. Salivary SARS-CoV-2 IgG MIA validation

Both bead batches (8-million and 20-million coupling scale) were used for threshold setting and were subsequently validated as described below. Using the 20-million coupling scale, we had introduced an assay standard and were able to validate the MFI-based result and the result based on standardized arbitrary units (AU).

The 8-million bead batch resulted in 98.6% sensitivity (142/144 samples collected >14 days post COVID-19 symptoms onset classified correctly as positive) and 99.2% specificity (263/265 pre-COVID-19 era classified correctly as negative) at a cutoff of six and passed validation with very similar performance. The proportion of pre-COVID-19 era and COVID-19 study participant samples combined that tested negative and did not pass the total IgG concentration requirement was 22.7% (Error! Reference source not found.). For the 20-million batch a cutoff of 9 (MFI) and 10 (AU) were chosen to maximize assay specificity. The resulting sensitivity (96.7%) was marginally lower in the threshold setting sample set but almost all (80/81 [98.8%]; AU-based classification) or all (81/81 [100%]; MFI-based classification) of the COVID-19 study participant samples collected >14 days post symptoms onset were classified correctly as positive during assay validation. There was very little difference between relying on the standardized classification based on AUs compared to using net MFI.

### 3.4. Algorithms to classify prior SARS-CoV-2 natural infection

We evaluated SARS-CoV-2 anti-N IgG classification accuracy by tabulating the anti-N IgG result (Algorithm 4) among samples classified as SARS-CoV-2 IgG positive (anti-N/RBD/S IgG positive, algorithm 7). This sequence resulted in a sensitivity of 94.1% (95% confidence interval [CI]: 90.0%, 96.9%) and specificity of 99.2% (95% CI: 97.7%, 99.8%) for detection of anti-N IgG positives, which indicates prior exposure to or natural infection with SARS-CoV-2.

### 3.5. SARS-CoV-2 IgG MIA precision

Contrived high and low positive controls and a negative control near the limit of detection (LLOQ) were tested in replicate over several days to assess within-plate and between-plate assay precision. The overall result (sum[S/Co]) and the signal (MFI and AU) to individual antigens were assessed. The coefficient of variation (CV) of the result between replicates within batch was under 5%. Replicates of the high and low control also resulted in less than 5% variation from the mean for individual antigens. The CV of the of the LLOQ control for individual antigens was between 0% and 33%. The assay precision between plates/days was around 10% for the sum of S/Co to N/RBD/S and ranged between 7% and 13% for the high and low controls and between 11% to 32% for the LLOQ control when assessing each antigen signal individually (**Table S4-S7**).

The average sum of S/Co of 10 pre-COVID-19 era saliva samples was 1.0 (range: 0.3-1.6); all tested negative and serial dilutions of a contrived sample resulted in 93% to 110% recovery and 117% recovery near the lower limit of quantitation (**Tables S8-S9**). The standard error of replicates of contrived saliva samples near the cutoff was 2%. All samples contrived to result either 25% or 50% above or below the cutoff were classified correctly (**Table S10**). The assay precision using MFI for analysis was nearly identical (not shown).

### 3.6. Calibration of salivary SARS-CoV-2 IgG MIA to U.S. SARS-CoV-2 serology standard

Using the U.S. national serology standard as calibrator, we estimated the SARS-CoV-2 IgG concentration equivalent of our assay cutoff as ∼0.1 SARS-CoV-2 IgG BAU/mL (see **Figure 4**). The range for detection of SARS-CoV-2 binding IgG in oral fluid was 0.002 to ∼4.0 BAU/mL using our MIA. The anti-S and anti-N IgG concentration in BAU/mL relative to the WHO SARS-CoV-2 serological standard (1,000 BAU/mL) is 764 BAU/mL and 681 BAU/mL, respectively, for the US SARS-CoV-2 serological standard and 371 BAU/mL and 330 BAU/mL, respectively, for our undiluted in-house serological standard. The cutoffs for individual N, RBD and S antigens in the assay range between 0.04 and 0.13 BAU/mL (see Figure 4). Note that the final cutoff values (sum of S/Co to N, RBD and S) is higher than the average of the individual cutoffs to maximize assay specificity.

**Figure 4.**
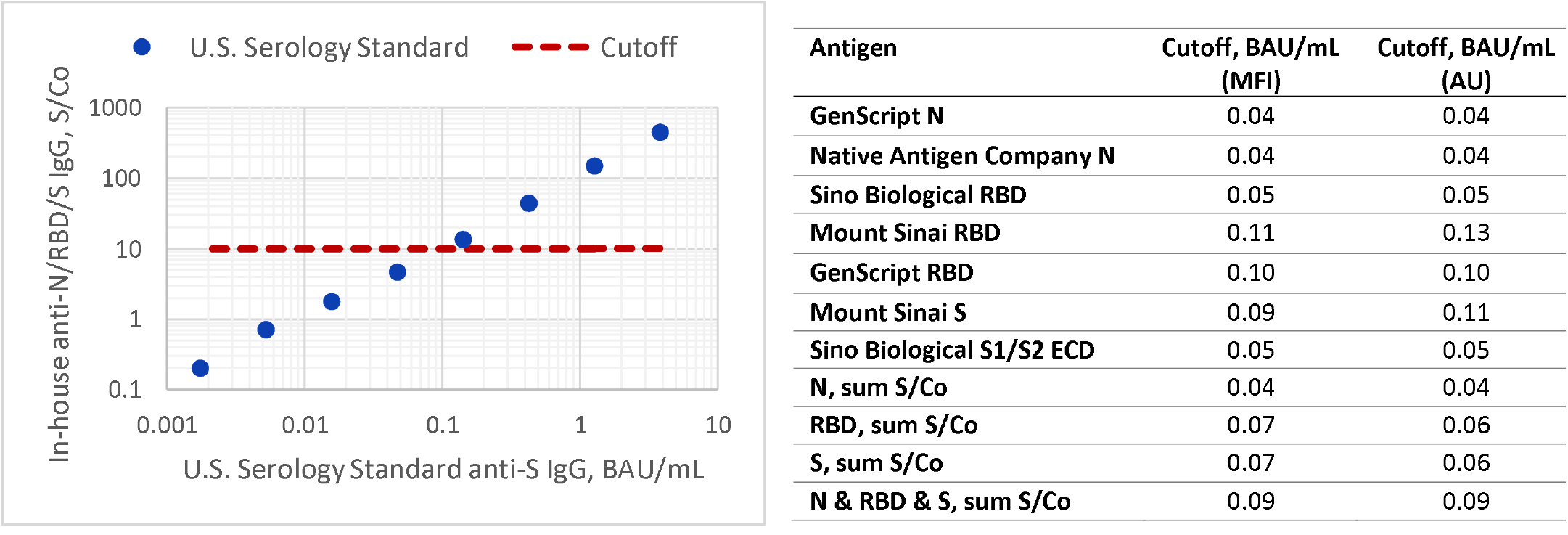
Calibration of oral fluid sum of signal to cutoff (S/Co) for N, RBD, and S to the US SARS-CoV-2 serology standard binding anti-S IgG binding antibody units (BAU) per mL. Note. N: nucleocapsid; RBD: receptor binding domain; S: spike; ECD: ectodomain.

### 3.7. Correlation of neutralizing antibody with blood- and oral fluid-based SARS-CoV-2 IgG response

Plasma samples with neutralizing antibody and IgG binding data to RBD, spike and N antigens were available for 30 of the study day 28 samples. Matching saliva samples were used to estimate correlations between blood and salivary IgG measurements; indeterminates were excluded. IgG to N, RBD, and spike in blood correlated with the corresponding measure in saliva (N: rho=0.76, RBD, rho=0.83, spike: rho=0.82; all *p*<0.001). Salivary IgG levels to RBD (rho=0.78; *p*<0.001) and spike (rho=0.79; *p*<0.001) correlated equally well or slightly better with plasma neutralizing antibody AUC than plasma IgG levels (RBD: rho=0.79, spike: rho=0.76), whereas IgG to N (rho= 0.68) in saliva correlated less strongly with neutralizing antibody than IgG to N in plasma (rho=0.76) (see **Figure 5**).

**Figure 5.**
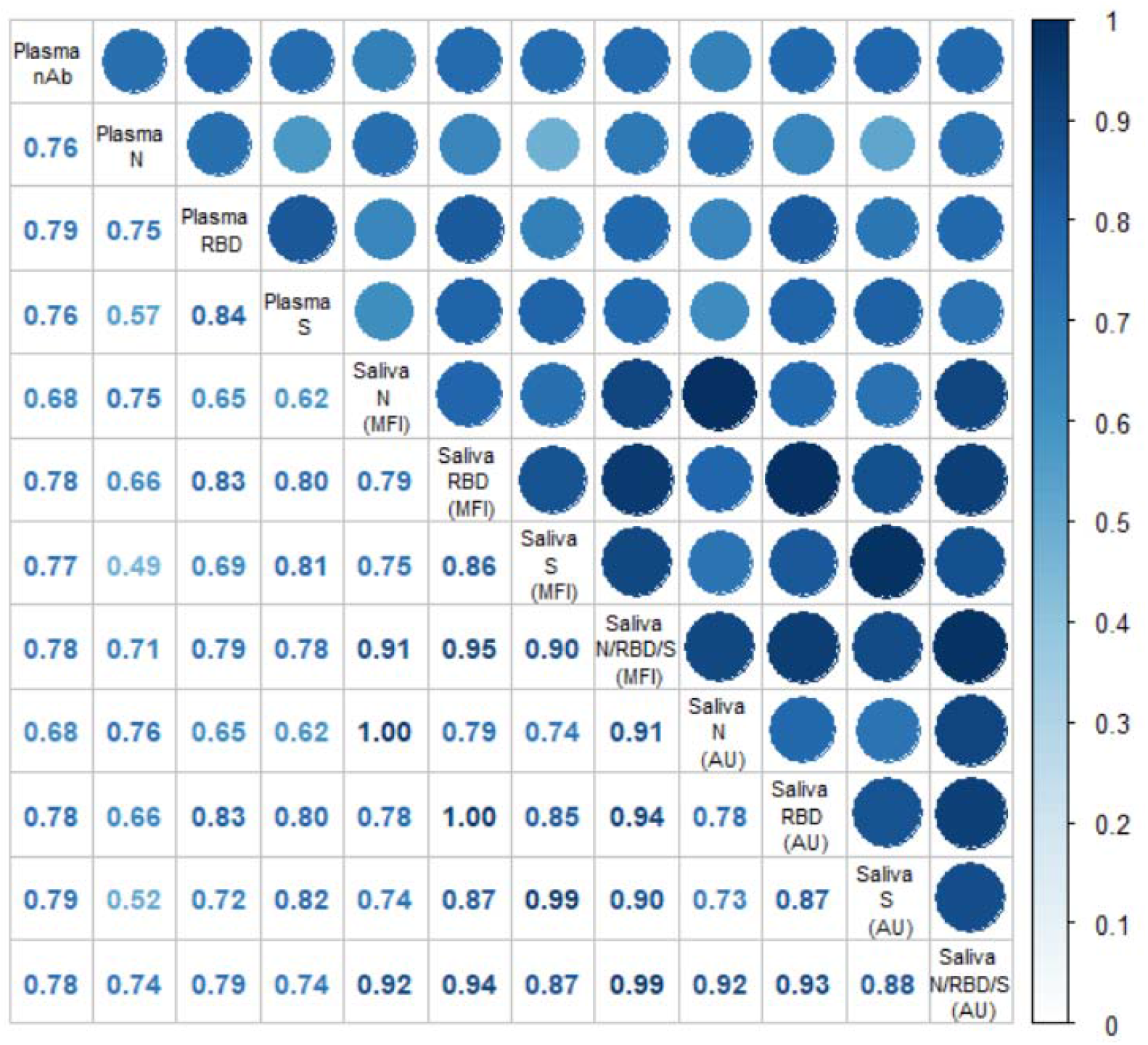
Correlation between plasma SARS-CoV-2 nAb, plasma SARS-CoV-2 IgG ELISA and salivary SARS-CoV-2 IgG MIA for n=30 matched samples. Note: Plasma anti-N, anti-RBD and anti-S IgG area under the curve (AUC) determined by ELISA, plasma neutralizing antibody AUC (nAb) determined by neutralizing antibody assay. Saliva N, RBD or S AU/MFI: sum of multiplex assay signal to cutoff of anti-N IgG, anti-RBD or anti-S IgG or the sum of S/Co to all antigens (N/RBD/S) measured either in mean fluorescence intensity (MFI) or arbitrary units (AU).

### 3.8. Salivary antibody kinetics post SARS-CoV-2 infection

In accordance with the improved assay performance upon removing samples with indeterminate classification, this concept also refines SARS-CoV-2 IgG kinetics after COVID-19 symptoms onset. **Figure 6A** shows a Loess regression with all samples, whereas in **Figure 6B** indeterminate samples were excluded. The exclusion of potentially false-negative samples due to insufficient total salivary IgG leads to a steeper average rise in SARS-CoV-2 IgG and most individuals reach maximum SARS-CoV-2 IgG levels earlier, similar to average time to seroconversion (9 days +/-3) observed in blood samples^25^. Most individuals “sero”converted for saliva SARS-CoV-2 IgG around 9 days post symptoms onset and had detectable IgG responses to the three antigen types used in the assay (N, RBD and spike, **Figure 7**). After 14 days post-symptom onset 202/211 samples (96%) tested positive for anti-N IgG, 193/199 (97%) tested positive for anti-RBD IgG, and 181/193 (94%) tested positive for anti-S IgG. The average time to seroconversion was slightly faster for nucleocapsid-binding IgG (8.0 days) than for RBD-binding (8.4 days) or spike-binding IgG (10.1 days, **Figure 7**). However, comparing the salivary anti-N, RBD and S kinetics to each other rather than in reference to when they cross the cutoff we did not see any difference in the time to reaching a plateau between the antigens (see **Figure S3**). Saliva samples average a concentration of 0.8 anti-N IgG and 0.5 anti-spike IgG BAU/mL >2 weeks post symptoms onset with maximum concentrations of 3.4 anti-N IgG and 4.0 anti-S IgG BAU/mL.

**Figure 6.**
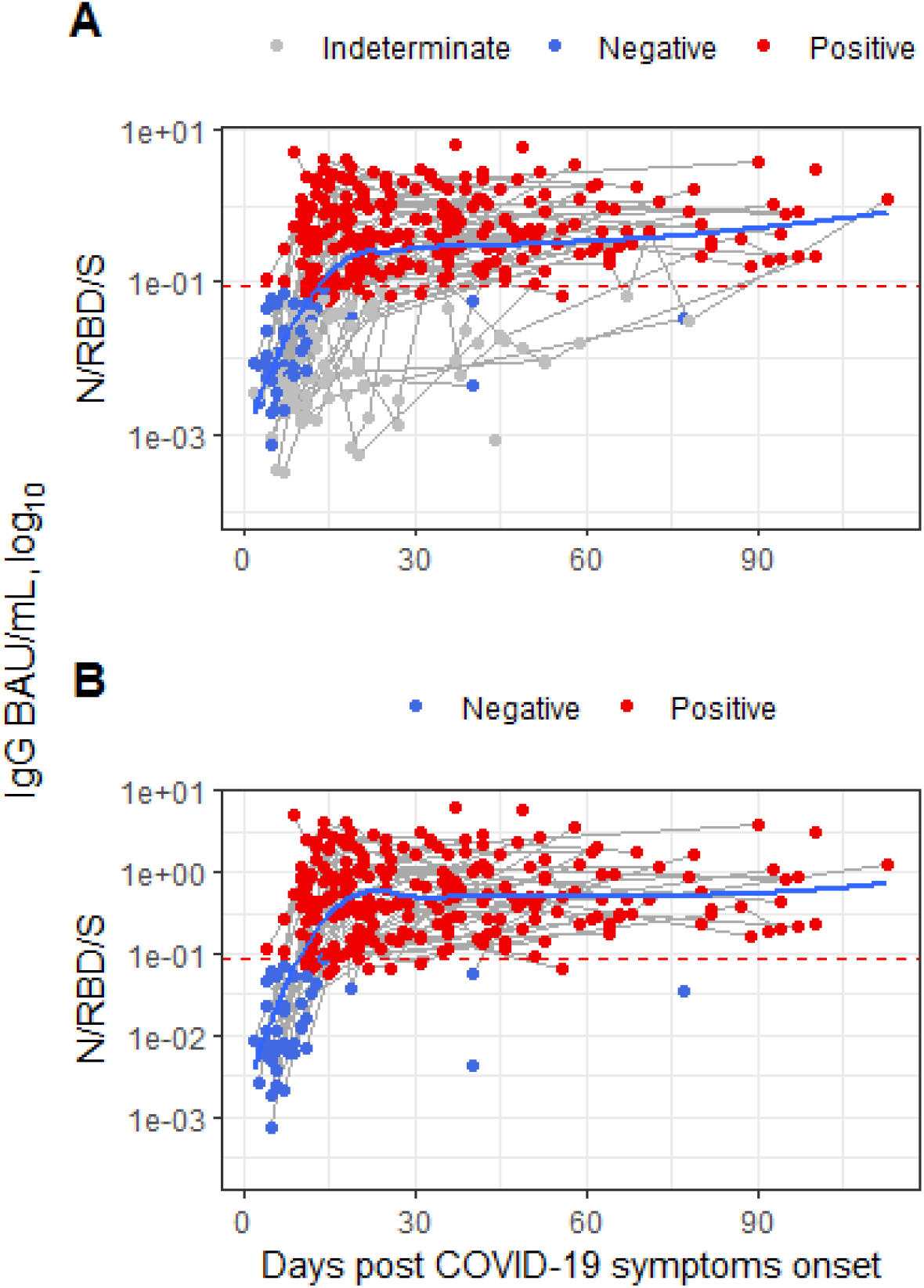
SARS-CoV-2 IgG kinetics post infection in saliva. **A**. Samples classified as SARS-CoV-2 positive, negative and indeterminate contributed to Loess regression (n=375). **B**. Samples with insufficient total IgG that tested negative (grey points) were excluded, leading to more naturally plausible IgG signal progression, similar to antibody kinetics observed in blood (n=288). Grey lines connect longitudinal samples from the same participant. **Note**. Y-axis scale reflects a calibration of our in-house sum of anti-N/RBD/S IgG signal to cutoff to the U.S. SARS-CoV-2 serological standard anti-S IgG BAU/mL.

**Figure 7.**
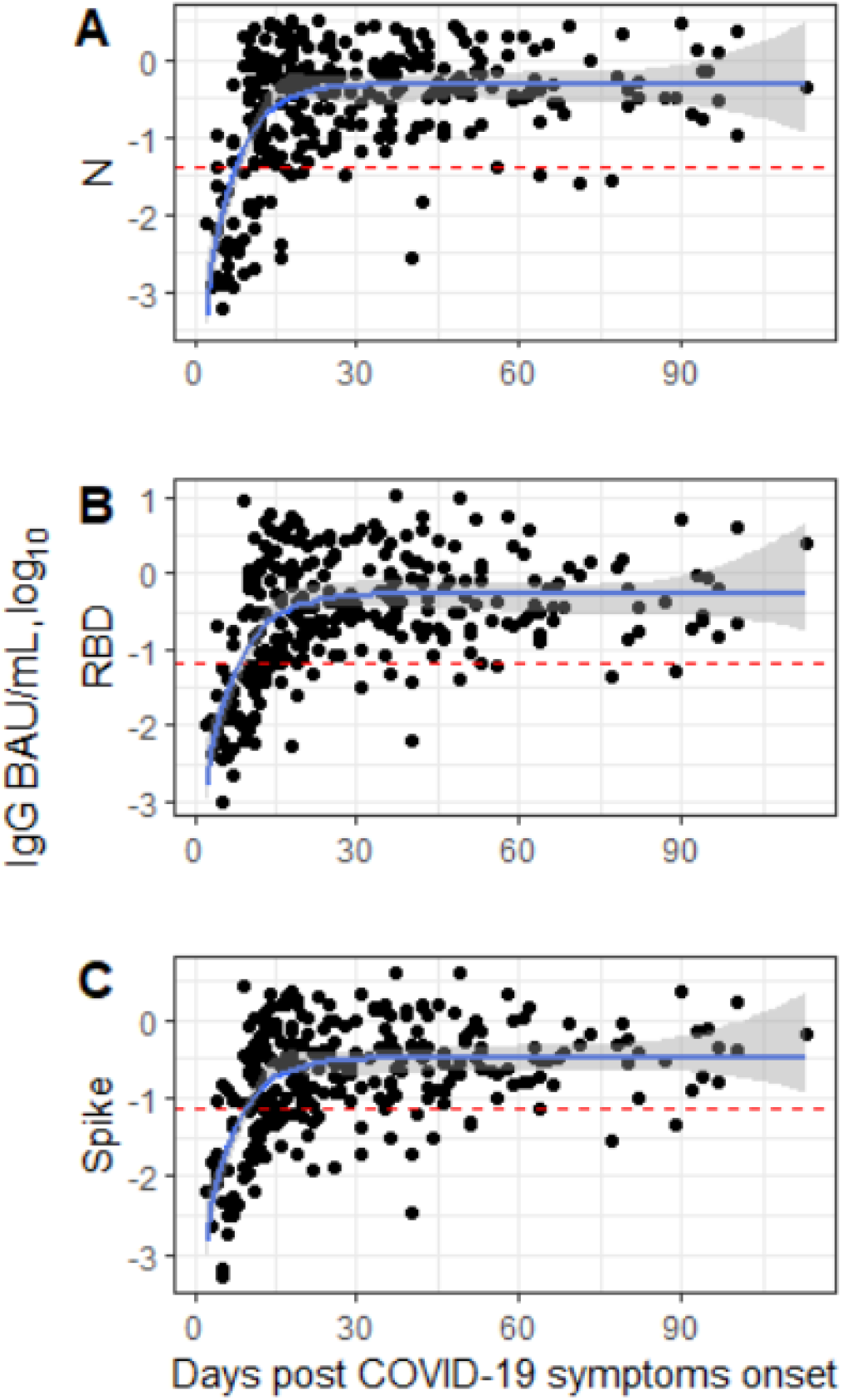
Anti-N, RBD and Spike IgG (BAU/mL) kinetics in saliva collected by COVID-19 study participants over time. **A**. Anti-nucleaocapsid (N) IgG. **B**. Anti-receptor-binding domain (RBD) IgG. **C**. Anti-spike (S) IgG. Blue line represents a monomolecular model (exponential growth function) fitted to the log10-transformed IgG concentration. Red dotted lines represent the cutoff.

## 4. DISCUSSION

We established a SARS-CoV-2 antibody assay for saliva with performance characteristics comparable to the best performing blood-based antibody tests.^26^ We accomplished this by characterizing and addressing the assay limitations systematically. First, we set out to characterize the assay performance for each component, which led us to conclude that relying on individual antigens of the assay would at best classify ∼80% of positive samples correctly and ∼98% of negative samples. Next, we explored combining IgG signals to multiple antigens, which improved the assay performance somewhat (85% sensitivity and 99% specificity). We then examined differences between samples that were classified as false-negatives and those that were classified correctly as positives, which showed that samples with low total IgG concentration were much more likely to produce false-negative results. This observation resulted in the establishment of a “quality control step”. The quality control step fails samples that tested negative for SARS-CoV-2 IgG and contain less than 15 μg/mL total IgG. This added QC step greatly improved the assay’s performance, which was, with some additional fine-tuning of the cutoff, comparable to the best blood-based SARS-CoV-2 antibody assays with >98% sensitivity and >99% specificity. The assay performance was nearly identical at two different bead coupling scales.

However, the QC step also leads to sample loss (indeterminate classification), because a significant proportion of saliva samples cannot be classified with the recommendation that the participant collects a new sample. If this assay were based on blood, this would be an unacceptable request, particularly for children, however, we have found that study participants do not mind providing a second sample if their saliva sample could not be classified as long as they were provided with a second collection swab. We have also found that the number of samples with indeterminate classification (i.e., with insufficient total IgG) can be greatly reduced when the sample collection procedure is explained well and at least for the first time supervised either by video-call or in-person.

The salivary total IgG concentration is influenced by the sample collection device, the type of saliva that is being collected (passive drool, parotid, sublingual, gingival crevicular fluid, etc.), collection technique, and duration of sample collection. Other factors include time of day, time since last meal and drink, participant age and gum health. Saliva sample collection devices, clear instructions for sample collection and sample processing can be standardized and even diurnal effects could be offset by instructing participants to collect samples, e.g., 30 minutes after their usual breakfast. However, other factors like age (which affects total salivary antibody concentration ranges) and gum health likely need to be controlled for through additional measurements, ideally within the same multiplex assay. If most of these factors were either eliminated or normalizers existed that allow to adjust for the between-sample differences, then the proportion of the indeterminate class would likely shrink greatly.

We showed that salivary and plasma anti-N, RBD and S IgG correlate highly between the two sample types and found that salivary SARS-CoV-2 anti-RBD and anti-S IgG binding estimates correlate equally well with neutralizing titers as corresponding blood-based estimates. Other groups have developed and characterized assays to measure salivary SARS-CoV-2 antibodies primarily using ELISA with mixed success.^11,12,27^ Typically, the sensitivity of oral fluid assays is at least 10% lower than the sensitivity of serum or plasma assays if cutoffs are optimized such that >99% specificity is maintained and researchers conclude that testing salivary antibody testing may complement traditional serology without being a stand-alone alternative to blood-based testing.^28–30^ To overcome the challenge associated with low total IgG concentrations in saliva compared to blood some have resorted to concentrating saliva, either with or without additional accounting for salivary total IgG, which resulted in increased assay sensitivity but reduced specificity.^31^ Despite the lower assay performance most groups found that blood SARS-CoV-2 IgG levels correlate well with salivary IgG levels^32^, however, such findings are often based on very small sample sizes.^31,33,34^ Studies with larger sample sizes typically report less robust correlations between saliva and blood antibody levels.^30,35^

In the Ambulatory COVID-19 Study cohort nearly all participants had seroconverted by saliva IgG to SARS-CoV-2 two weeks post COVID-19 symptoms onset and SARS-CoV-2 IgG levels were stable for the duration of the study. We did not observe a significant difference in time to seroconversion and time to plateau between anti-N, anti-RBD and anti-S IgG in saliva. To our knowledge, this is the first study to report salivary anti-N and anti-spike IgG concentrations in oral fluid post infection that were calibrated to the international SARS-CoV-2 antibody standard in BAU/mL, which seem approximately 200-500-fold lower than those in serum or plasma. We measured a median anti-S IgG concentration of 0.3 BAU/mL (mean 0.5 BAU/mL) in saliva collected between 14 days and 90 days post infection, whereas others reported a median serum anti-spike concentration of 154 anti-spike BAU/mL post infection (using the Roche Elecsys anti-S assay).^36^ Our in-house assay cutoff of 0.09 anti-spike IgG BAU/mL for saliva is ∼200-fold lower than the Ortho Vitros spike IgG cutoff for serum/plasma cutoff that has been established at 17.8 BAU/mL^37^, ∼375-fold lower than the Diasorin Trimeric S IgG cutoff of 33.8 BAU/mL but, compared to the Elecsys anti-SARS-CoV-2 S cutoff of 0.8 anti-S BAU/mL, our in-house cutoff is only ∼10-fold lower. This demonstrates that even though efforts have been made to harmonize quantitative SARS-CoV-2 humoral immune responses between laboratories, cutoffs for binary classification of antibody status and also quantitative assay ranges between tests that have been calibrated to the WHO international or the U.S. serological SARS-CoV-2 standard still vary. For example, cutoffs for anti-spike IgG positivity in serum or plasma range from 0.8 BAU/mL to 33.8 BAU/mL between just five commercial anti-RBD/S assays used in a comparative “head-to-head” study and means of anti-spike concentrations post vaccination were reported to vary between ∼70 BAU/mL and ∼1500 BAU/mL using the same set of samples.^38,39^ Despite absolute concentration discrepancies, most serological SARS-CoV-2 total Ig and IgG binding assays showed good quantitative correlations (rho>0.8).^38^ The Diasorin Trimeric S IgG assay and the Elecsys anti-SARS-CoV-2 S assay are among the best performing antibody assays for blood^26^, however, their respective cutoffs (0.8 anti-spike BAU/mL for Roche Elecsys and 17.8 BAU/mL for Ortho Vitros) vary by factor of >22; although the absolute difference is small (<20 BAU/mL). Panels comprising SARS-CoV-2 antibody negative and low, medium and high titer samples that have been quantified and characterized with the most commonly used serologic assays would be a useful reference for better comparison between reported testing data. Continued standardization and calibration of assays for saliva but also for serum and plasma will be important to confirm absolute concentration differences between the different specimen types and to harmonize cutoffs for sample classification.

Salivary SARS-CoV-2 IgG testing will be useful to determine the robustness of antibody responses post vaccination^40^, differences in antibody response between the different SARS-CoV-2 vaccine types, and could be a tool to assess whether and when vaccine boosters may be needed due to waning immunity^41^ at the individual level. Several groups have reported on salivary anti-S/RBD responses post vaccination, which elicits IgG responses that are at least 10-fold higher^42^ than post natural infection and are therefore more easily detected in saliva.^40,43^ Additionally, salivary SARS-CoV-2 anti-N IgG testing may continue to be a useful estimate of exposure or infection among both vaccinated and naïve individuals. Saliva as a non-invasive specimen that can deliver SARS-CoV-2 antibody prevalence estimates with high accuracy is particularly attractive in studies with repeated sampling time points and when phlebotomy is either impractical, too costly or not an option, for example in large serosurveys, remote locations, when young children are participating or in elderly cohorts.^30,35,44^ Salivary secretory IgA responses to SARS-CoV-2 as a surrogate of mucosal immunity^45–47^ may predict protection from (re-) infection and new intra-nasal vaccines that elicit a stronger mucosal antibody response than intramuscular vaccines might protect better from infection than currently available intra-muscular vaccines.^48,49^ Salivary multiplex assays will be an important tool to assess the role of preexisting and cross-priming mucosal antibodies that may exhibit cross-reactivity with other endemic coronaviruses (229E, HKU1, NL63 and OC43) and their potential role in COVID-19 prevention and progression.^50^

## Data Availability

Deidentified research data produced in the present study are available upon reasonable request to the authors.

