## Supplemental Material for "Methodological approaches to optimize multiplex oral fluid SARS-CoV-2 IgG assay performance and correlation with serologic and neutralizing antibody responses"

**Supplement**

**Table S 1**. Multiplex assay antigens, controls, and secondary antibody.

| No. | Vendor / Producing Lab | Antigen/Antibody | Cat. No. |
| --- | --- | --- | --- |
| 1 | Native Antigen Company | Nucleoprotein | REC31851 |
| 2 | GenScript | Nucleoprotein | Z03480 |
| 3 | Sino Biological | Spike S1 RBD | 40592-V08H |
| 4 | Mt. Sinai | RBD | n/a |
| 5 | GenScript | RBD | Z03483 |
| 6 | Mt. Sinai | Spike | n/a |
| 7 | Sino Biological | S1+S2 ECD | 40589-V08B1 |
| 8* | Native Antigen Company | S1 | REC31806 |
| 9 | Native Antigen Company | SARS 2002 N | REC31744 |
| 10 | Sino Biological | SARS 2002 RBD | 40150-V08B2 |
| 11 | Native Antigen Company | MERS S1 | REC31760 |
| 12 | Sino Biological | hCoV NL63 S1+S2 ECD | 40604-V08B |
| 13 | Sino Biological | hCoV 229E S1+S2 ECD | 40605-V08B |
| 14 | Sino Biological | hCoV OC43 HE | 40603-V08H |
| 15 | Sino Biological | hCoV HKU1 S1 | 40021-V08H |
| 16 | Sino Biological | RSV RSS2 | 40037-V08B |
| 17 | Sino Biological | RSV A2 | 11049-V08B |
| 18 | Jackson ImmunoResearch | anti-IgG | 109-005-098 |
| 19 | Jackson ImmunoResearch | anti-IgA | 109-005-011 |
| 20 | Jackson ImmunoResearch | anti-IgM | 109-005-129 |
| 21 | Pierce | BSA | 23225 |
| Detect Ab | Jackson ImmunoResearch | anti-human IgG-PE | 109-115-098 |

Note: *This antigen was excluded due to poor performance. Ab=antibody

**Table S 2**. Sensitivity and specificity of individual SARS and SARS-CoV-2 antigens in the multiplex test

| No. | Antigen | N | Sensitivity | Specificity |
| --- | --- | --- | --- | --- |
| 1 | GenScript N | 536 | 133/174 (76.4%) | 356/362 (98.3%) |
| 2 | Native Antigen N | 536 | 136/174 (78.2%) | 354/362 (97.8%) |
| 3 | Sino Biological RBD | 536 | 146/174 (83.9%) | 355/362 (98.1%) |
| 4 | Mount Sinai RBD | 536 | 121/174 (69.5%) | 354/362 (97.8%) |
| 5 | GenScript RBD | 536 | 136/174 (78.2%) | 359/362 (99.2%) |
| 6 | Mount Sinai S | 536 | 102/174 (58.6%) | 359/362 (99.2%) |
| 7 | Sino Biological S1/S2 ECD | 536 | 136/174 (78.2%) | 356/362 (98.3%) |
| 8 | Native Antigen S1 | 536 | 2/174 (1.2%) | 359/362 (99.2%) |
| 9 | SARS 2002 N | 536 | 133/174 (76.4%) | 356/362 (98.3%) |

**Table S 3**. Multiplex assay performance using algorithms that combine IgG signals to multiple antigens

|  |  | **COVID-19 patient samples** | | | **pre-COVID-19 samples** |
| --- | --- | --- | --- | --- | --- |
|  |  | Days after COVID-19 symptoms onset | | |  |
|  |  | 1-7 days | 8-14 days | >14 days |  |
|  | n | 41 | 78 | 174 | 362 |
| **Algorithm** | **Contributing antigens** | **Sensitivity** | | | **Specificity** |
| IgG signal above cutoff for multiple antigens (crosshairs) | | | | | |
| **1** | ≥1 N & ≥1 RBD or S | 4.9% | 43.6% | 78.2% | 99.4% |
| **2** | ≥2 RBD or S | 9.8% | 46.2% | 81.0% | 99.2% |
| **3** | ≥1 N & ≥1 RBD or S  or ≥2 RBD or S | 9.8% | 51.3% | 83.3% | 98.6% |
| Sum of IgG S/Co to multiple antigens | | | | | |
| **4** | N antigens (n=2) | 9.8% | 47.4% | 79.3% | 97.8% |
| **5** | RBD antigens (n=3) | 9.8% | 44.9% | 81.0% | 98.6% |
| **6** | S antigens (n=2) | 14.6% | 48.7% | 75.9% | 98.9% |
| **7** | N & RBD & S antigens (n=7) | 9.8% | 53.8% | 84.5% | 98.6% |
| Sum of IgG S/Co normalized with total IgG concentration | | | | | |
| **8** | N & RBD & S S/CO divided tIgG | 2.6% | 20.0% | 43.8% | 98.6% |

Note. Assay performance assessed at 8-million beads coupling scale. S/Co: signal to cutoff; N: nucleocapsid; RBD: receptor binding domain; S: spike; tIgG: total salivary IgG in μg/mL. Samples are classified using either binary outcomes (algorithms 1-3) or continuous outcome, i.e. the sum of signal to cutoff (algorithms 4-8). Cutoffs for individual antigens were defined as the mean + 3 SD of pre-pandemic negative saliva samples.

a) 8-Million bead batch (MFI)

Sensitivity/Specificity (%)

Excluded (%)

| Assay cutoff | Sensitivity | Specificity | Indetermiante |
| --- | --- | --- | --- |
| 4 | 99.3333% | 96.6165% | 21.5094% |
| 5 | 98.6301% | 98.4906% | 22.3062% |
| 6 | 98.6111% | 99.2453% | 22.6843% |
| 7 | 98.5714% | 99.2453% | 23.4405% |
| 8 | 98.5612% | 99.6226% | 23.6295% |
| 9 | 97.8417% | 99.6226% | 23.6295% |
| 10 | 95.6204% | 99.6226% | 24.0076% |

b) 20-Million bead batch (MFI)

Sensitivity/Specificity (%)

Excluded (%)

| Assay cutoff | Sensitivity | Specificity | Indetermiante |
| --- | --- | --- | --- |
| 4 | 98.4733% | 94.9772% | 20.0965% |
| 5 | 97.6562% | 97.2603% | 20.8065% |
| 6 | 97.619% | 98.6239% | 21.4516% |
| 7 | 96.748% | 98.6239% | 21.8094% |
| 8 | 96.7213% | 99.0826% | 22.0779% |
| 9 | 96.7213% | 100% | 22.2222% |
| 10 | 95.8678% | 100% | 22.8099% |

c) 20-Million bead batch (AU)

Sensitivity/Specificity (%)

Excluded (%)

| Assay cutoff | Sensitivity | Specificity | Indetermiante |
| --- | --- | --- | --- |
| 4 | 99.2366% | 95.8904% | 19.9085% |
| 5 | 97.6562% | 96.8037% | 20.595% |
| 6 | 97.6378% | 98.1651% | 21.0526% |
| 7 | 97.6% | 99.0826% | 21.5103% |
| 8 | 97.561% | 99.0826% | 21.789% |
| 9 | 96.7213% | 99.5413% | 22.0183% |
| 10 | 96.7213% | 100% | 22.0183% |

**Figure S1**. Assay performance at different cutoffs to classify pre-pandemic presumed negative saliva samples and saliva collected >14 days post COVID-19 symptoms onset using bead batches coupled at a) 8-million beads coupling scale; b) and c) 20-million beads coupling scale. For a) and b) the median fluorescence intensity (MFI) was used to determine presence of SARS-CoV-2 IgG; c) standardized arbitrary units (AU) were used to determine presence of SARS-CoV-2 IgG.

**Assay Precision**

Table S4. Within day/batch precision of combined index

| Sample Type | Replicates, n | sum (S/Co) | SD | CV |
| --- | --- | --- | --- | --- |
| High control | 10 | 78.7 | 2.6 | 3% |
| Low control | 10 | 25.1 | 0.8 | 3% |
| LLOQ | 2 | 0.1 | 0.0 | 2% |

Table S5. Within day/batch precision of individual antigen components

| High control | Replicates, n | AU | SD | CV |
| --- | --- | --- | --- | --- |
| GenScript N | 10 | 5021 | 174 | 3% |
| Native Antigen N | 10 | 4357 | 159 | 4% |
| Sino Biological RBD | 10 | 2044 | 68 | 3% |
| Mt. Sinai RBD | 10 | 1515 | 45 | 3% |
| GenScript RBD | 10 | 2423 | 80 | 3% |
| Mt. Sinai S | 10 | 1906 | 70 | 4% |
| Sino Biological S | 10 | 1427 | 38 | 3% |
| Low control |  |  |  |  |
| GenScript N | 10 | 1709 | 54 | 3% |
| Native Antigen N | 10 | 1472 | 54 | 4% |
| Sino Biological RBD | 10 | 581 | 18 | 3% |
| Mt. Sinai RBD | 10 | 430 | 14 | 3% |
| GenScript RBD | 10 | 732 | 26 | 4% |
| Mt. Sinai S | 10 | 530 | 14 | 3% |
| Sino Biological S | 10 | 431 | 9 | 2% |
| LLOQ |  |  |  |  |
| GenScript N | 2 | 5.1 | 0.0 | 0% |
| Native Antigen N | 2 | 4.8 | 0.3 | 7% |
| Sino Biological RBD | 2 | 5.1 | 0.2 | 3% |
| Mt. Sinai RBD | 2 | 3.2 | 1.1 | 33% |
| GenScript RBD | 2 | 4.7 | 0.2 | 5% |
| Mt. Sinai S | 2 | 5.1 | 0.1 | 3% |
| Sino Biological S | 2 | 4.8 | 0.0 | 0% |

Table S6. Between day/batch precision – combined index

| Sample Type | Batch | Replicates n | sum (S/Co) | SD | CV |
| --- | --- | --- | --- | --- | --- |
| High control | 1-10 | 40 (4 per batch) | 77.7 | 6.6 | 9% |
| Low control | 1-10 | 40 (4 per batch) | 24.4 | 2.0 | 8% |
| LLOQ | 1-10 | 20 (2 per batch) | 0.1 | 0.0 | 11% |

Table S7. Between day/batch precision – individual components

| High control | Replicates n | Mean | SD | CV |
| --- | --- | --- | --- | --- |
| GenScript N | 40 | 4842 | 537 | 11% |
| Native Antigen N | 40 | 4366 | 403 | 9% |
| Sino Biological RBD | 40 | 2086 | 193 | 9% |
| Mt. Sinai RBD | 40 | 1493 | 138 | 9% |
| GenScript RBD | 40 | 2405 | 196 | 8% |
| Mt. Sinai S | 40 | 1917 | 169 | 9% |
| Sino Biological S | 40 | 1350 | 99 | 7% |
| Low control |  |  |  |  |
| GenScript N | 40 | 1639 | 130 | 8% |
| Native Antigen N | 40 | 1412 | 114 | 8% |
| Sino Biological RBD | 40 | 599 | 75 | 12% |
| Mt. Sinai RBD | 40 | 418 | 44 | 10% |
| GenScript RBD | 40 | 717 | 82 | 11% |
| Mt. Sinai S | 40 | 536 | 68 | 13% |
| Sino Biological S | 40 | 401 | 26 | 7% |
| LLOQ |  |  |  |  |
| GenScript N | 20 | 4.8 | 0.5 | 11% |
| Native Antigen N | 20 | 4.5 | 0.5 | 12% |
| Sino Biological RBD | 20 | 4.4 | 0.6 | 15% |
| Mt. Sinai RBD | 20 | 4.0 | 1.3 | 32% |
| GenScript RBD | 20 | 4.4 | 0.5 | 12% |
| Mt. Sinai S | 20 | 4.3 | 0.5 | 13% |
| Sino Biological S | 20 | 4.3 | 0.6 | 13% |

Table S8. Assessment of 10 saliva samples collected prior to the COVID-19 pandemic

| Pre-COVID-19 era sample | sum (S/Co) |
| --- | --- |
| Sample 1 | 0.3 |
| Sample 2 | 1.6 |
| Sample 3 | 1.1 |
| Sample 4 | 0.9 |
| Sample 5 | 1.5 |
| Sample 6 | 0.6 |
| Sample 7 | 0.4 |
| Sample 8 | 1.5 |
| Sample 9 | 0.4 |
| Sample 10 | 1.3 |

Table S9. Linearity and recovery of spiked serum sample (arbitrary units)

| Linearity | Replicates, n | Spiked  sum (S/Co) | Measured  sum (S/Co) / SD | | Recovered, % |
| --- | --- | --- | --- | --- | --- |
| Sample 1 | 2 | 35.8 | 33.3 | 0.01 | 93% |
| Sample 2 | 2 | 11.9 | 11.1 | 0.29 | 93% |
| Sample 3 | 2 | 4.0 | 4.2 | 0.36 | 104% |
| Sample 4 | 2 | 1.3 | 1.5 | 0.03 | 110% |
| Sample 5 | 2 | 0.4 | 0.5 | 0.00 | 106% |
| Sample 6 | 2 | 0.1 | 0.2 | 0.00 | 117% |

Table S10. Assessment of samples near the assay cutoff (combined index)

| Sample Type | Replicates, n | Mean sum (S/Co) | SD | CV |
| --- | --- | --- | --- | --- |
| Close to cutoff (10 au) | 30 | 9.6 | 0.2 | 2% |
| 50% above cutoff (15 au) | 20 | 14.1 | 0.3 | 2% |
| 50% below cutoff (5 au) | 20 | 4.9 | 0.1 | 2% |
| 25% above cutoff (7.5 au) | 20 | 6.9 | 0.2 | 2% |
| 25% below cutoff (12.5 au) | 20 | 12.7 | 0.2 | 2% |

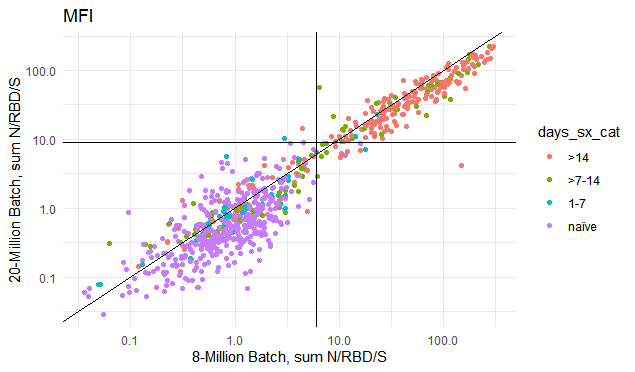

**Figure S2. Between-batch comparison of the anti-N, RBD and S IgG IgG signal (sum signal/cutoff).** Antigens were coupled at 8-Million and 20-million beads scale to assess between-batch variability. Horizontal and vertical lines show the assay cutoff for each batch.

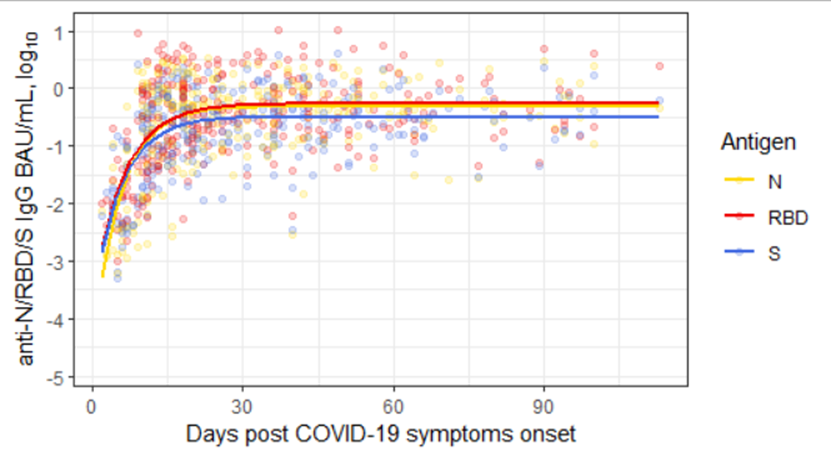

**Figure S3.** Anti-N, RBD and Spike IgG (BAU/mL) kinetics in saliva collected by COVID-19 patients over time. Solid lines represent a monomolecular model (exponential growth function) fitted to the log10-transformed IgG concentration.
